# Comparing lateral flow testing with a rapid RT-PCR method for SARS-CoV-2 detection in the UK

**DOI:** 10.1101/2021.10.08.21264742

**Authors:** Andrew Taylor, Ronan Calvez, Mark Atkins, Colin G Fink

## Abstract

In late 2019, SARS-CoV-2 emerged in the Wuhan province of China. Rapid global spread led to the Covid-19 pandemic. Rapid and accurate detection of SARS-CoV-2 has become a vitally important tool in controlling the spread of the virus. Lateral flow devices (LFDs) offer the potential advantage of speed and on-site testing. The sensitivity of these devices compared to the gold standard RT-PCR has been questioned. We compared the performance of the Innova lateral flow kit, recommended by the UK government, with our rapid in-house RT-PCR protocol using stored positive patient samples. The LFD device was found to be 6,000-10,000 times less sensitive than RT-PCR for the detection of SARS-CoV-2. Overall, the LFD detected 46.2% of the positives detected by RT-PCR. 50% of the LFD results were observed to be weak positives, only visible after careful examination by experienced laboratory staff. At lower viral loads, such as 10,000-100,000 RNA copies/ml, the LFD detected 22.2% of positives. In addition, two strong positives (3 and 1.5 million RNA copies/ml) were not detected by the LFD. The argument for use of LFD kits, despite their lack of sensitivity, is that they detect infectious virus and hence contagious individuals. At present, there is a lack of scientific evidence supporting this claim. The LFD used in the UK fails to identify individuals with considerable viral loads and has been subject to a class I recall by the US FDA but is still approved and recommended for use by the UK government. We believe that using LFD testing for assessing SARS-CoV-2 infection risk is a strategy which has risks that outweigh any benefits.

## Introduction

In late 2019, a novel coronavirus emerged in the Wuhan province of China. This virus was designated severe acute respiratory syndrome coronavirus-2 (SARS-CoV-2) and rapid global spread lead to the Covid-19 pandemic. The symptoms of infection vary between individuals but the most common symptoms are fever, dry cough, headache, fatigue, anosmia and diarrhoea^1,2^. As of 13^th^ June 2021, there have been 175 million global cases with 3.79 million deaths^3^. The UK has been badly affected with 4.55 million cases and the highest official death toll in Europe (>127, 000). The devastating nature of this pandemic facilitated the need for high-throughput and accurate diagnostic methods. RT-PCR protocols for the detection of SARS-CoV-2 RNA were published very early in the pandemic with the first publication detailing probe-based RT-PCR assays targeting E, N and RdRp genes^4^. Shortly after this, the US CDC published three assays (all probe-based RT-PCR assays) which target three separate regions of the N gene^5^. These assays were given the names N1, N2 and N3 although the N3 assay was later removed as it was deemed unreliable. Many of these published assays have been developed as commercial kits and are used globally for the detection of SARS-CoV-2 RNA. We developed a highly sensitive, rapid, UKAS accredited in-house RT-PCR assay utilising the N1 assay published by the CDC^6^. RNA is prepared using a rapid heat treatment method, removing the need for time-consuming and expensive nucleic acid extraction kits^6^.

RT-PCR has the advantage of being the most sensitive diagnostic technique available and is regarded as the gold standard for SARS-CoV-2 detection. As the pandemic progressed, the possibility of using rapid, point of care devices such as lateral flow devices (LFDs), has been further investigated. Such kits target viral proteins and thus claim to detect live virus and hence infectious individuals^7^. They claim to offer the advantage that results can be obtained in as little as 30 minutes reportedly without the need for skilled staff or laboratory processing. However, there have been questions about the sensitivity of such kits, particularly when compared to RT-PCR^8-11^.

We retrospectively compared the UK government supplied Innova LFD with RT-PCR using a range of samples that had previously tested as SARS-CoV-2 positive using our diagnostic PCR service. Using digital droplet PCR to calibrate clinical samples, we were able to directly compare PCR (in terms of RNA copies/ml) with the LFD device currently recommended for use in the UK for assessing infection risk of individuals in schools, universities, hospitals and for the general public.

## Methods

### Patient samples

Initially, 62 stored swab samples were selected for testing. This comprised 52 known PCR positive samples (original Ct value 18.9-36.5) and 10 negatives. Samples were previously tested as part of our routine diagnostic service and had been stored at -20°C for up to 3 months. Samples consisted of either dry swabs which were resuspended in 500μl of 0.1% Igepal CA-630 (Sigma Aldrich) or swabs in UTM. Following the emergence of the delta variant eight additional samples, which were confirmed delta variants, were also tested.

### RT-PCR

Samples were prepared for RT-PCR using a rapid heat-treatment method^6^. This involves heating samples to 95°C for 5 minutes and cooling to 4°C. 40μl of sample was heat treated and 10μl used for PCR (25μl reaction volume). RT-PCR was then carried out in triplicate using Fast 1-Step RT-qPCR (Meridian Bioscience)^6^. To further explore comparative sensitivity of RT-PCR and LFD, a strongly positive sample (sample 29) was quantified using droplet digital (dd) PCR (QX200, Bio-Rad) and serially diluted from 2.2 million to 220 RNA copies/ml in 0.1% Igepal CA-630. Triplicate PCR was carried out as described above.

The lower limit of detection (LLoD) of this method was determined by Probit regression analysis (MedCalc, MedCalc Software Ltd) using a diluted RNA extract which was quantified by ddPCR.

### Lateral flow

Lateral flow kits used for SARS-CoV-2 testing in the UK (Innova rapid antigen test, manufactured by Xiamen Biotime Biotechnology) were directly compared to the RT-PCR method. Instructions provided with the kit state that 2 drops of extraction buffer should be applied to an LFD strip. Therefore, the volume of 2 drops of buffer from an extraction tube was measured using a calibrated pipette and found to be 50μl. To standardise all LFD tests, this volume of sample was added to each LFD using a pipette. Frozen patient samples were thawed and 40μl mixed with 160μl of LFD extraction buffer by vortexing. From this, 50μl was applied to LFD strips in triplicate. Using this method, the same equivalent volume of patient sample (10μl) was tested on each LFD compared to each single RT-PCR reaction allowing direct comparison of the two procedures under standardised conditions. Kits were used following the manufacturers’ instructions and results scored independently by 2 experienced laboratory staff in standardised light conditions. The LLoD was determined as above using Probit regression analysis. The dilution series of sample 29 (described above) was also tested in triplicate on LFDs.

### Statistical analysis

Probit regression (MedCalc) was used to determine the LLoD (with 95% confidence interval) of both methods. These were calculated using either the Ct values or the viral load (in RNA copies/mL) as measured by ddPCR. The correlation between viral load and LFD result was calculated in Excel by first converting the LFD results into a score of 0-3 reflecting the number of replicates with a positive result. The Pearson correlation value (r) was then calculated.

### Ethical statement

This study was performed at Micropathology Ltd (University of Warwick Science Park, Coventry, UK) in April – June 2021. Patient samples were anonymised and were not considered Human Subjects Research due to the quality improvement and public health intent of the work. This study was reviewed and approved by the Micropathology Ltd Ethics Committee Review Board composed of Professor Sheila Crispin (MA, VetMB, DVA, DVOphthal, DipEVCO, FRCVS), Professor Christopher Dowson (BSc, PhD), Rt Hon Countess of Mar, Most Rev Dr Gordon Mursell (MA, Hon DD) and William NH Taylor (BTech)). No additional consent was necessary.

## Results

### Sample set

Approximately equal numbers of samples from male and female patients were tested (Table S1). The age of patients ranged from 4 to 92 and the mean Ct value from the original PCR results was 30. No samples with an original Ct value >37 (<3000 RNA copies/ml) were tested.

### Comparing RT-PCR with LFD

Summarising the data for the 62 samples tested, all ten of the previous PCR negatives were negative on all three replicates of PCR and LFD (Fig. 1). Of the 52 previously positive samples, 51 were positive in all 3 PCR replicates and one sample was positive in 2 out of 3 replicates. Of these 52, only 24 (46.2%) returned positive LFD results in at least 2 out of 3 replicates. Considering viral load, samples with a viral load of > 1 million copies/ml resulted in an 85.6% detection rate on LFD. Surprisingly, two samples in this range (3 million and 1.5 million copies per ml) were negative in all 3 LFD replicates. Preliminary sequencing data (not shown) suggests that both of these samples contain the Alpha variant, the dominant variant at the time the samples were taken. In the range of 100,000 – 1 million copies per ml, 46.7% of the samples were positive in at least 2 of the 3 replicate LFD tests. The sensitivity of LFD was very limited in the range of 10,000-10000 RNA copies/ml (22.2%) and even lower at <10,000 copies/ml (15.4%). A statistically significant correlation between Ct value (or log viral load) and LFD result was observed (r = 0.59, P < 0.001).

**Fig. 1:**
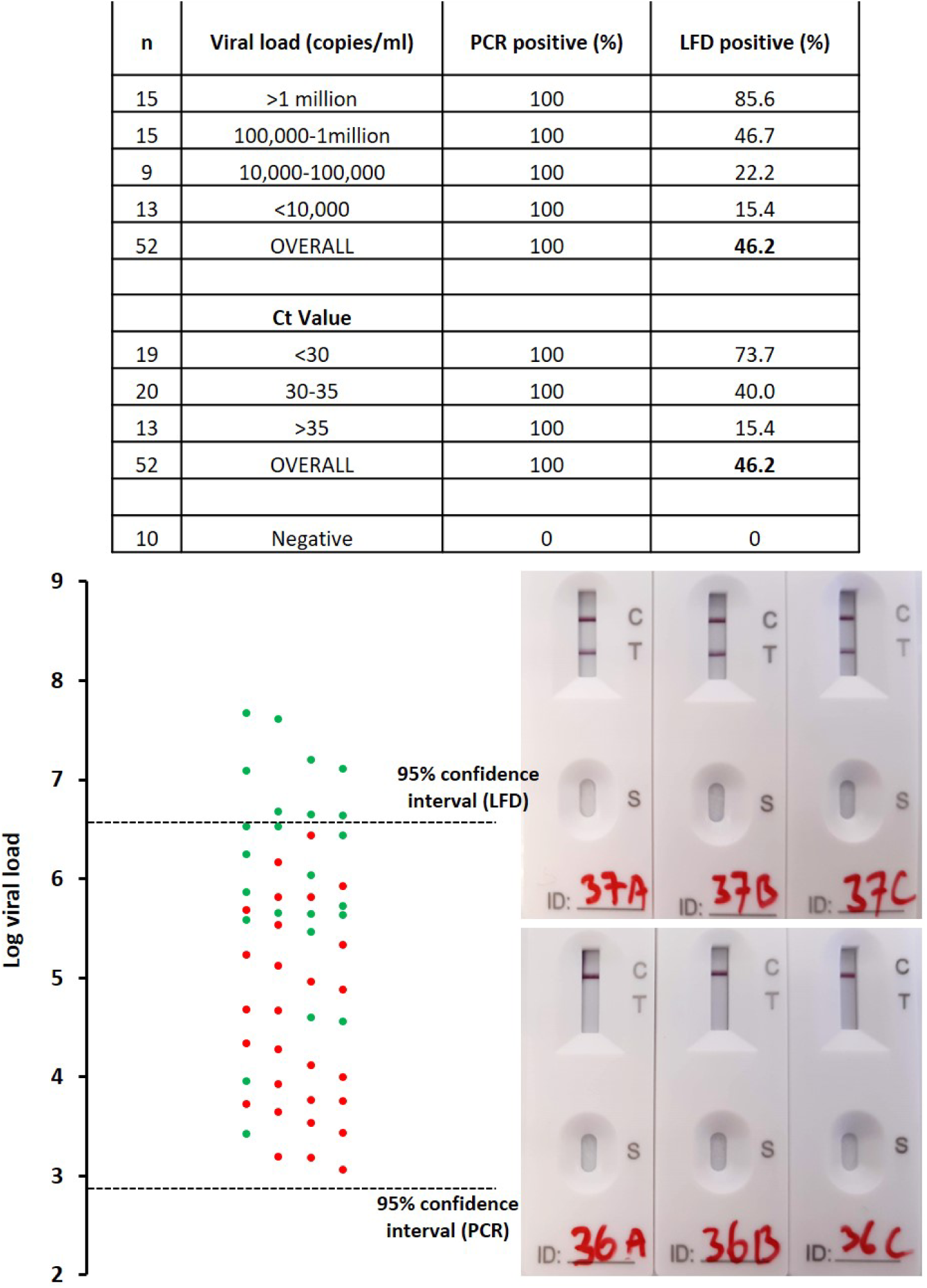
Comparing LFD with RT-PCR. Samples which were positive in at least 2 out of the 3 replicates were classified as positive. A green data point represents a sample where at least 2 out of 3 replicate tests were positive. A red data point indicates a sample where 0 or 1 of the 3 replicates were positive. Of the 52 positive samples tested, 51 were positive in all 3 RT-PCR replicates. One sample was positive in 2 out of 3 RT-PCR replicates. Photographs of the LFD device show examples of positive (top) and negative (bottom) test results. The 95% confidence interval was calculated using a Probit regression analysis (Fig. 2).

### LOD of LFD and RT-PCR

The Probit analysis (Fig. 2a) revealed that the LOD of the RT-PCR assay, when run as a duplex assay with an internal control, is 743 copies/ml (95% confidence interval 508-1,559 RNA copies/ml; P < 0.0001). The sensitivity can be further improved to 681 copies/ml by doubling the reaction volumes (95% confidence interval 540-822 RNA copies/ml: P < 0.0001; data not shown). Using the data summarised in Fig. 1, we performed a Probit analysis (Fig. 2b) to determine the LLoD of the LFD devices (expressed in terms of RNA copies/ml for direct comparison with RT-PCR). The LLoD was found to be 4.35 million copies/ml (95% confidence interval 2.75-12.0 million copies/ml, P < 0.0001), suggesting that the LFD devices were ∼6,000 times less sensitive than RT-PCR.

**Fig. 2:**
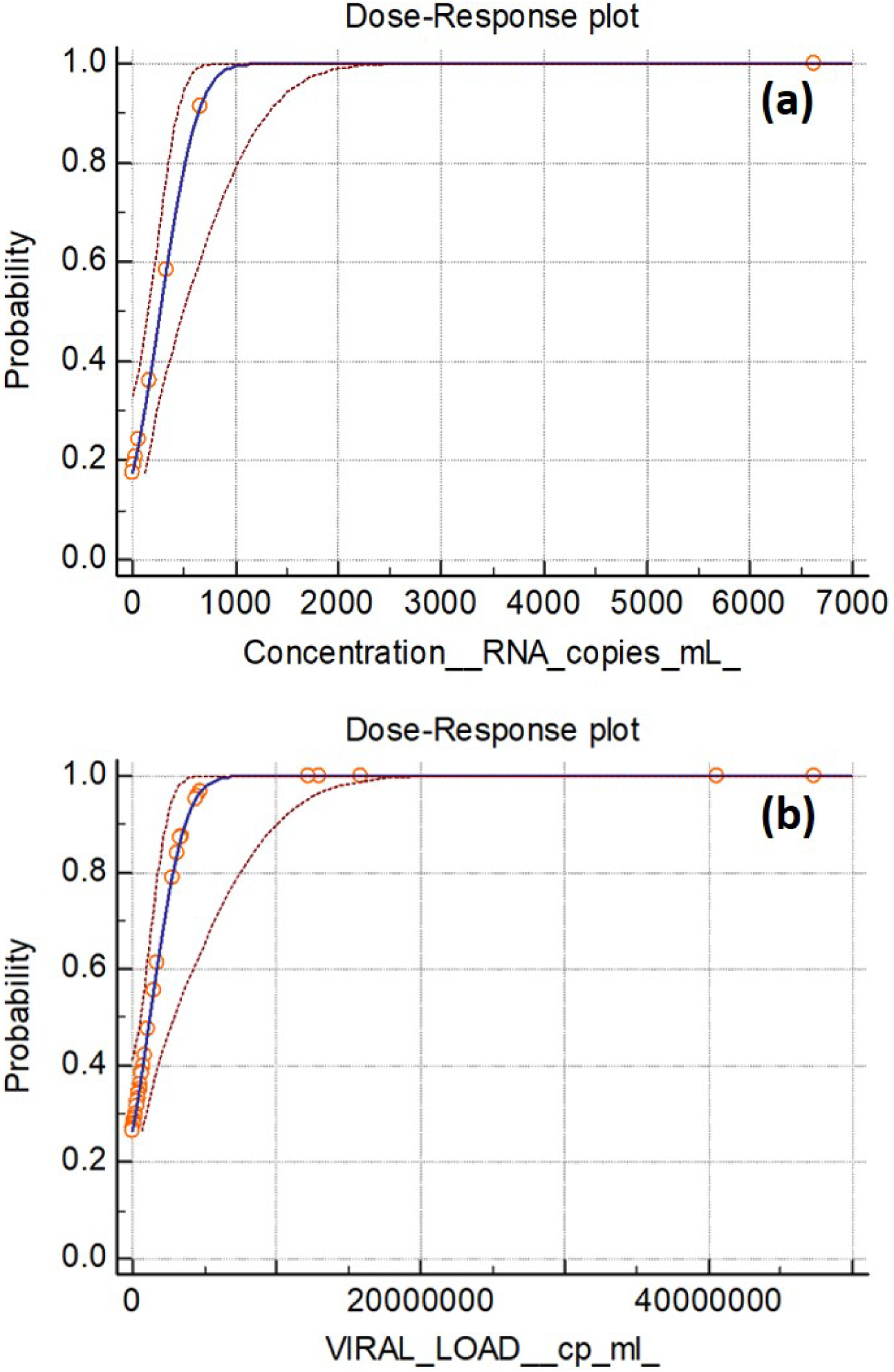
Evaluation of the lower limit of detection of the RT-PCR assay (a) and the Innova rapid antigen LFD (b) expressed as RNA copies per ml. Probit regression was used to calculate the LLoD of the LFDs using the data summarised in Fig. 1 compared to RT-PCR when multiplexed with an internal control (potato virus Y, PVY).

To further explore the sensitivity of LFD devices, a dilution series of a strongly positive sample (22 million copies/ml) was tested. A strong correlation between viral load (digital PCR) and Ct value (RT-PCR) was observed (r=-0.998, P < 0.001, Fig. 3). All LFD tests were negative below 2.2 million copies/ml although PCR was successful down to 220 copies/ml. This indicates that the PCR test used is at least 10,000 times more sensitive than LFD, which is consistent with the data obtained above, using undiluted samples.

**Fig. 3:**
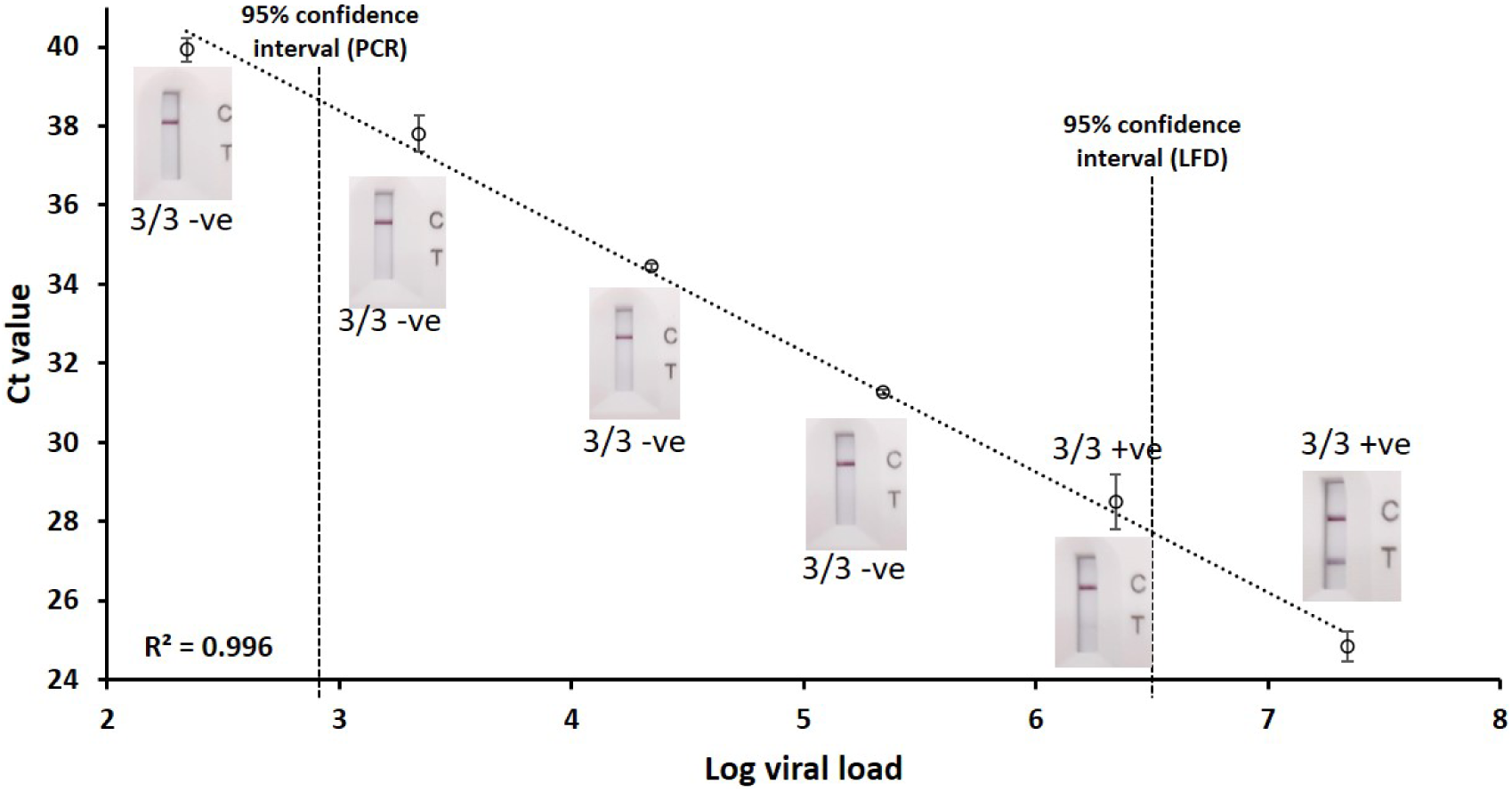
Correlating Ct value (RT-PCR) with viral load (digital PCR). Results of LFD tests are shown for each dilution. The 95% confidence intervals were calculated based on the Probit analysis (Fig. 2).

Ct values of all SARS-CoV-2-positive samples tested as part of a diagnostic service from 1^st^ – 31^st^ Jan 2021 were analysed (Fig. 4). The mean Ct value was 32.7 (range 19.5 – 41.9). As part of this diagnostics service, all samples with a Ct value greater than 39 were re-extracted and the PCR repeated for confirmation. Based on the 95% confidence interval obtained from the Probit analysis, LFD testing would only have detected 104 out of 501 positives (20.8%).

**Fig. 4:**
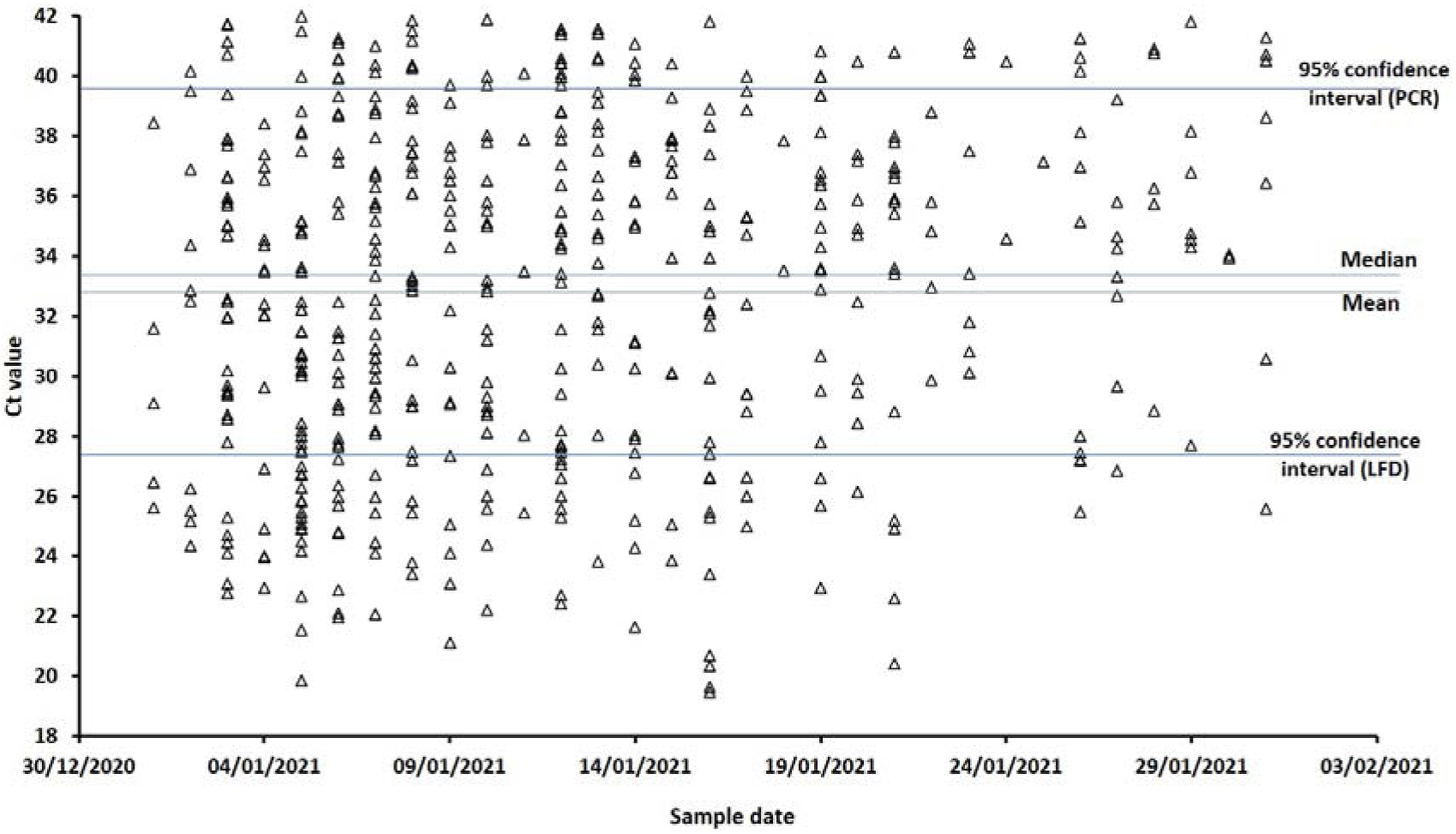
Ct values obtained from positive patient samples tested from 1^st^– 31^st^ January 2021. 95% confidence intervals were calculated using Probit analyses.

### Detection of the delta variant

Due to the rapid spread of the delta variant in the UK, eight samples which were confirmed to contain the delta variant were tested by LFD and PCR. The results were comparable with the data described above (Fig. 1). Samples with a Ct value of >30 were not detected by LFD (Table 1). Samples with a Ct value of 26-30 were detected but scored as very weak.

**Table 1:**
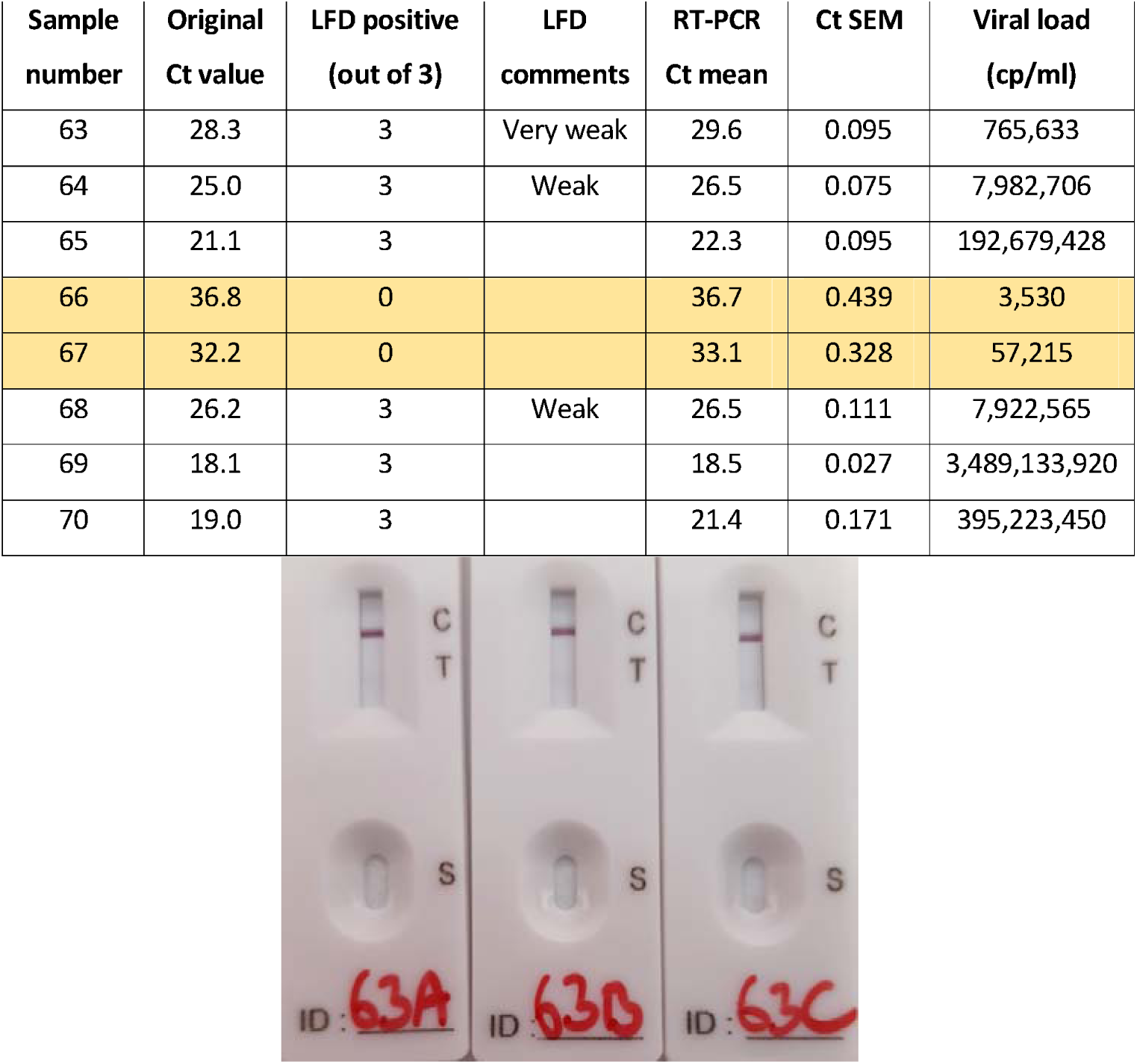
Testing the Innova LFD kit against delta variants of SARS-CoV-2. The image illustrates a ‘weak positive’ LFD result. SEM is the standard error of the mean based on 3 replicates.

## Discussion

The UK government has been promoting the use of Innova LFD tests for rapid screening and home testing to identify individuals infected with SARS-CoV-2. Data published by the Department of Health and Social Care suggests that such tests have a high level of clinical specificity and thus a low rate of false positives^12^. The figures quoted are up to 100% specificity but the sensitivity of LFD tests is somewhat overlooked. Government data from multiple testing sites shows a sensitivity of 37.4 – 57.4% when compared to PCR (viral load of samples not specified). Our data is in agreement with these figures, using 52 positive patient samples we found a sensitivity of 46.2%. In reality, this means that LFD devices detect less than half of the positives detected by PCR. However, when considering the range of positives detected in our laboratory in a calendar month, the sensitivity of LFD is likely to be much lower – a figure of 20.8% was calculated based on a 95% confidence interval. An argument supporting the lack of sensitivity of LFD is that they are sufficient to detect ‘live’ virus as shown by some cell culture experiments^7,13,14^. The theory is that they should identify patients who are contagious. It is not clear whether all of the positive patients tested by our laboratory would be contagious and present a transmission risk. However, it is clear that the virus was present within their respiratory tract as determined by the presence of RNA. Whilst a proportion of these patients may be carrying residual/non-functional RNA or RNA coated in antibodies, it is likely that some patients with a low viral load are at an early stage of infection, before symptom presentation. Data published by Public Health England clearly shows that samples with Ct values of 32.5 can contain live virus (1.2 PFU/ml) whilst detection by LFD is very poor in samples above Ct 28.5 (40 PFU/ml)^15^. In a separate study, also by Public Health England, it was shown that viable SARS-CoV-2 virus can be cultured from samples with Ct values >35^16^. Therefore, there is no general consensus as to the relationship between viral load and infectiousness. We diluted a strong positive sample which was positive on PCR and LFD (assumed to be live virus). When diluted 10-fold, the LFD result was still positive but was negative when diluted 100-fold (220,000 copies/ml). RT-PCR remained positive down to 220 copies/ml. Any correlation between detection of live virus and LFD positive is unproven and LFD devices simply lack sensitivity to detect SARS-CoV-2 reproducibly.

It has been noted by others that Ct values only provide a guide on viral load and there is no consensus on a Ct value which corresponds to infectiousness^7,17^. Therefore, we used digital PCR to correlate Ct values with viral loads. Government LFD testing data used samples with unspecified viral loads^12^. Previous data tested samples ranging from <1000 - >1 million copies/ml^15^. However, it is not clear how these viral loads were calculated. The authors quote a Ct value of 25.5 corresponding to 100,000 copies/ml although, using our in-house assay, a Ct value of 25.5 was equivalent to 17 million copies/ml.

The LFD tested here failed to detect SARS-CoV-2 in samples with viral loads of 3 million and 1.5 million copies/ml. This observation was also noted during large scale community testing in Liverpool where LFD sensitivity was only 67% in samples with a Ct value <25^18^. Furthermore, the studies cited above used cell culture experiments to test for live virus and infer infectivity of a patient on this basis^7,13,14^. This data should be interpreted with caution; epidemiological data from Switzerland showed that outbreaks occurred in areas where the first three cases had viral loads of <100,000 copies/ml^19^. Such cases would be missed by LFD testing and therefore transmission of the virus could follow. A patient tested at our laboratory was initially positive for SARS-CoV-2 during routine screening (Ct 38). He was put into isolation and started developing Covid symptoms two days later with concurrent increase in viral load. This shows the critical importance of detecting low levels of SARS-CoV-2 RNA in order to isolate patients. This initial sample would have been undetected using an LFD.

The overall sensitivity of the LFD device, based on the samples tested here, was 46.2%. However, it should be noted that 50% of the positive LFD tests (12/24) produced a very faint positive result (see image in Table 1) across all replicates. These were scored as positive by two experienced laboratory staff but may have been scored as negative by a member of the general public. In addition to this, the samples tested were known positives, providing a perfect sample for the LFD. The validity of self-administered tests and thus sample quality remains a concern, data published by the UK government showed that LFD sensitivity was drastically reduced from 79.2% to 57.5% when LFD tests were performed by members of the public compared to a trained individual^15^. Stories in the British media suggested that false positive results can be obtained using fruit juice^20^, something which we confirmed in our laboratory. In addition, the positive control does not confirm antigen detection or sample quality. This is of particular concern for self-administered tests.

The justification for using LFD devices is that they are rapid, cheap and can produce a result in around 30 minutes whilst bypassing the requirements for rigorous laboratory standards. Our streamlined laboratory PCR procedure is able to produce results in as little as 90 minutes from sample receipt without the need for formal nucleic acid extraction allowing us to offer same day results and our laboratory has been able to upscale to a capacity of 4000 tests a day. Whilst the cost of an individual LFD device is relatively low, the actual costs to the UK tax-payers are much larger. A recent audit found that of the 691 million test kits distributed, results had only been reported from 96 million (14%)^21^. There is no evidence of any cost-benefit analysis for LFD testing in the UK.

We conclude that the LFD device used in the UK has limited merit for reducing SARS-CoV-2 infection in the community as it is 6000 – 10000 times less sensitive than PCR. A negative result may provide a false sense of security and should be interpreted with extreme caution. It has been suggested that LFD devices could be used to test individuals 7 days after a positive PCR test to confirm that their viral load is low enough to allow safe release from quarantine^7,22^. However, an ongoing transmission risk may remain and a PCR test would still be more appropriate, both individually and for epidemiological purposes. We believe that the use of LFD testing for SARS-CoV-2 diagnostics in the UK (and globally) is a strategy which has risks that outweigh any potential benefits. Based on the data presented here, it is no surprise that the US FDA has issued a Class I recall for the Innova kit, citing concerns over the performance of the device^23^. Innova has been issued with a warning letter by the FDA relating to misleading data in the validation of the kit. Despite this, the device remains approved for use in the UK (by the Medicines and Healthcare products Regulatory Agency).

## Data Availability

The data produced in the present study is present in the manuscript or is available upon reasonable request to the authors.

## Supporting Information

**Table S1:** Details of patient samples used for this study.

