## Supplementary material for "Comparing lateral flow testing with a rapid RT-PCR method for SARS-CoV-2 detection in the UK": Table S1

**Table S1**: Details of patient samples used for this study.

| **Gender** |  |
| --- | --- |
| Male | 32 |
| Female | 30 |
| **Age** |  |
| Mean | 38.8 |
| Median | 34.5 |
| Range | 4-92 |
| **Original CT of positives** |  |
| Mean | 30.2 |
| Median | 30.7 |
| Range | 18.9 - 36.5 |
